# The British Thoracic Society survey of rehabilitation to support recovery of the Post Covid-19 population

**DOI:** 10.1101/2020.05.07.20094151

**Authors:** SJ Singh, A Barradell, N Greening, CE Bolton, G Jenkins, L Preston, JR Hurst

## Abstract

**Objectives:** Those discharged from hospital after treatment for Covid-19 are likely to have significant and ongoing symptoms, functional impairment and psychological disturbances. There is an immediate need to develop a safe and efficient discharge process and recovery programme. Pulmonary rehabilitation is well placed to deliver a rehabilitation programme for this group but will most likely need to be adapted for the post Covid-19 population. The purpose of this survey was to rapidly identify the components of a post-Covid-19 rehabilitation assessment and elements of a successful rehabilitation programme that would be required to deliver a comprehensive service for those post Covid-19 to inform service delivery.

**Design:** A survey comprising a series of closed questions and a free text comments box allowing for a qualitative analysis.

**Setting:** Online survey.

**Participants:** British Thoracic Society members and multi-professional clinicians, across specialities were invited to take part.

**Results:** 1031 participants responded from a broad range of specialities over 6 days. There was overwhelming support for early post discharge from hospital phase of the recovery programme to advise patients about the management of fatigue (95% agreed/ strongly agreed), breathlessness (94%), and mood disturbances (including symptoms of anxiety and depression) 92%. At the 6-8-week time point an assessment was considered important, focusing on the assessment of a broad range of possible symptoms and the need to potentially return to work. Recommendations for the intervention described a holistic programme focusing on symptom management, return of function and return to employment. The free text comments added depth to the survey and the need ‘not to reinvent the wheel’ rather adapt well established (pulmonary rehabilitation) services to accommodate the needs of the post Covid-19 population.

**Conclusion:** The responses indicate the huge interest and the urgent need establish a programme to support and mitigate the long term impact of Covid-19.

**Strengths and limitations:**

- Large and comprehensive survey conducted to guide the provision of post Covid-19 rehabilitation.
- The survey provides clear recommendations for the provision of advice and support immediately upon discharge, and recommendations for a programme of holistic rehabilitation 6-8 weeks post discharge based upon the existing pulmonary rehabilitation model.
- The survey engaged a wide range of specialities and experiences managing Covid-19
- The opinions of patients and carers be sought in an additional survey

**Funding statement:** This research received no specific grant funding from any funding agency in the public, commercial or not-for-profit sector.

**Competing interests:** All authors have completed the Unified Competing Interests Form at http://www.icmie.org.coi_disclosure.pdf

Dr. Singh reports grants from Actegy, grants from Pfizer, outside the submitted work.

Dr. Jenkins reports grants from Astra Zeneca, grants from Biogen, personal fees from Boehringer Ingelheim, personal fees from Daewoong, personal fees from Galapagos, grants from Galecto, grants from GlaxoSmithKline, personal fees from Heptares, non-financial support from NuMedii, grants and personal fees from Pliant, personal fees from Promedior, non-financial support from Redx, personal fees from Roche, other from Action for Pulmonary Fibrosis, outside the submitted work.

**Data sharing statement:** No additional data are available.

## Background

The global coronavirus pandemic has already resulted in tens of thousands of people being admitted to hospital for acute medical management in the past few months, a proportion of whom will have had a prolonged stay on Intensive Care Units (ICU). Those discharged from ICU are likely to exhibit significant on-going symptoms notably dyspnoea, fatigue and cough, functional impairment and psychological disturbances[1–3]. The larger cohort of people discharged after ward based care are likely to experience similar if less severe problems.

Although we have limited Covid-19 data so far, literature from the SARS outbreak would suggest that there is a considerable impact upon the individual with reduced functional performance and health status even at 6 months post discharge compared to normal values[4,5].

There is a pressing need to develop a safe and efficient discharge process to support patients at home remotely and secondly to set up a mechanism to review these individuals early in the post discharge phase to facilitate restoration of pre-morbid function and holistic well-being and, for many, a successful return to maximal function and work.

Pulmonary rehabilitation teams routinely assess and manage the rehabilitation needs of patients with chronic respiratory disease often with multiple co-morbidities, including cardiovascular, mental health and metabolic diseases[6,7]. There is a strong evidence base demonstrating that a centre-based supervised out-patient programme of education and physical activity, collectively termed pulmonary rehabilitation positively impacts breathlessness, anxiety, depression, health status and exercise capacity. Pulmonary rehabilitation is an interdisciplinary intervention that integrates a broad group of health care professionals including but not limited to physiotherapists, nurses, dietitians, pharmacists, psychologists, physicians, occupational therapists, exercise physiologists and graduates of the programme. Pulmonary rehabilitation is recommended in all national and international guidelines for chronic obstructive pulmonary disease (COPD) and other long term respiratory conditions [8–10] and the most recent Cochrane review[11] suggested that no further randomised controlled trials were needed to establish the benefit of pulmonary rehabilitation in COPD. The provision of pulmonary rehabilitation is demonstrably successful in clinical practice outside the context of research studies, UK data from over 7000 cases has been collected and reported as part of the National Asthma and COPD Audit Programme (Pulmonary Rehabilitation)[12], Furthermore, participants frequently have multiple long-term conditions that do not compromise the outcome, the common comorbidities recorded in the chronic respiratory population mirror those that have been reported in the Covid-19 population of hypertension, diabetes and cardiovascular disease[13,14].

However, the rehabilitation needs of the post Covid-19 population are likely more diverse than those commonly observed in COPD and potentially with different objectives than those for chronic lung disease. Early data from Wuhan indicates that the mean age of people hospitalised with Covid-19 was 52·0 (45·0–58·0) years[13] compared to 69.0(9) years reported in for a conventional pulmonary rehabilitation programme[15], although data from the UK ISARIC registry of 16749 admissions indicates the median age is 72 (57-82) years[16], more typical of the pulmonary rehabilitation population. However, given the widespread nature of the pandemic there will be a substantial number of younger patients, especially those admitted to ICU, and in some of the post Covid-19 patients their pre-morbid state is likely to be quite different, many may not have preexisting lung disease, and likely different levels of employment, usual levels of activity and exercise behaviours. Furthermore, there is an indication the post Covid-19 population is likely to have significant psychological and cognitive impairment particularly if management involved a stay on ICU[17]. There is a small literature describing pulmonary rehabilitation interventions in the SARS population and ARDS with positive outcomes[18]. We postulated that whilst the core of pulmonary rehabilitation would in part meet the needs of the post Covid-19 patient, the programme would likely need to be adapted. The modifications would be at the point of assessment to broaden the scope to holistically assess the impact of Covid-19 and secondly to address the components of a comprehensive programme.

Therefore the purpose of this survey was to rapidly identify the additional components of a post-Covid-19 rehabilitation assessment, and elements of a successful rehabilitation programme that would be required to deliver a comprehensive service for those either discharged from hospital post Covid-19, or for those who we managed in the community but had marked ongoing symptoms that have prevented a full recovery.

## Methods

We conducted a survey of multi-professional clinicians. The survey was designed by team a with expertise in pulmonary and cardiac rehabilitation and the wider management of respiratory disease. The survey, supported by the British Thoracic Society (BTS), was predominately composed of closed questions, with a free text box at the end of the survey for additional comments. The survey was built by the team at the BTS using ClassApps software. The survey was tested by local teams experienced in rehabilitation prior to the wider launch.

The initial stages of the survey asked for basic demographic information from the participants to include age, gender, ethnicity, professional background, location of work and exposure to patients with, or recovering from Covid-19 (the full survey is available in the on-line supplement).

The purpose was to gain wide clinical consensus opinion as to what an effective, holistic rehabilitation intervention might comprise for patients recovering from Covid-19. The survey aimed to secure guidance for rehabilitation support provided in two phases:

- The initial discharge period (which may be to home, a step-down unit or a rehabilitation facility).
- A formal rehabilitation programme that would be offered 6-8 weeks post rehabilitation. This time period is based upon evidence accumulated by an ad-hoc task force formed by the ATS/ERS with a supporting document[19].

For these sections of the survey there was a statement and participants were invited to respond with the following five categories: ‘strongly disagree’, ‘disagree’, ‘neutral’, ‘agree’ or ‘strongly agree’. There was also the option to respond, ‘unable to comment’.

Upon completion of the survey there was an additional free text box for further comments. No questions were mandatory.

### Survey distribution

The survey was available to participants from 9^th^ April 2020 to 15^th^ April 2020. It was distributed to members of the BTS the societies’ e-newsletter and additionally promoted via the BTS Twitter account. A reminder email was sent to BTS members 6 days later and retweeted by society members and the BTS encouraging individuals to participate and share the survey with colleagues. The survey was not restricted to UK based health care professionals, although country of practice was noted on the survey.

Participation in the survey was voluntary and anonymous. Participants confirmed their willingness to engage in this research by accessing and completing the online survey. As the survey was directed towards health care professionals there was no patient or public involvement.

### Data analysis

Quantitative data were reported as counts and percentages for each category of the demographic and survey responses. At least 70% agreement on directionality (combining strongly agree and agree) was defined as the threshold for consensus.

Qualitative data were analysed using Thematic Analysis [20]. The text data was uploaded to NVivo Pro and then coded and grouped into themes to portray patterns within the data. The established themes were reviewed by the first author and the finalised themes were defined.

Ethical approval was not required for this survey from either the UK Health Research Authority or leading Research and Development Centre. Completion of the survey was an indication of willingness to participate and implied consent. We set no threshold for response over such a short period of time but were anticipating around 300 responses across a range of health care professionals to allow the questionnaire to be considered robust and representative of those in the field.

## Results

This report is based on data from 1031 respondents. A further 750 respondents only provided answers to the demographic questions on page 1 of the and therefore do not form part of this report. The majority of respondents were female (84%), the largest age group was 35-44 years (34%), followed by 45-54 years (27%) and 25-34 years (22%). A significant majority identified as white British (75%), with any other white background, white Irish and Indian representing 7%, 6% and 5% of the participants respectively. The largest group of respondents were from England (80%), with Scotland representing 6% of respondents, Wales 4%, Northern Ireland 3% and the remainder from the rest of Europe, Australasia and South America. Respiratory (32%) represented the largest known group by specialism, followed by Health Care of the Elderly (12%), Primary Care (7%) Acute Medicine (6%) and Sport & Exercise (5%) with smaller numbers from cardiology, general medicine, anaesthetics, psychiatry and psychology backgrounds. However, 34% recorded ‘Other’ backgrounds which included neurology (n=44), critical care (n=26) and musculo-skeletal (n=56)). Physiotherapists represented the largest group (71%), dietitians, nurses and consultant physicians represented 7%, 6% and 6% respectively, smaller numbers of occupational therapists, speech and language therapists, trainee physicians and health care assistants participated. The majority worked in either acute trusts (45%) or community hospitals/services (28%), with fewer responses from either primary care or private hospitals, 17% of ‘others’ were represented by the private (business) sector, universities, the community (excluding hospitals) and hospices. The final two profiling questions allowed for more than one response, in total there were 1420 respondents to whether or not they had experience in manging patients with Covid-19. 361 respondents had no experience, the remainder had experience on acute wards (n=332), Intensive Care Units (n=257), community (n=209) and step-down units (n=154). With respect to rehabilitation, 442 respondents had no experience, 216 in pulmonary rehabilitation, 208 in Health Care of the Elderly, 52 in cardiac rehabilitation and 202 in other forms of rehabilitation. Of the 1030 respondents 167 (16%) answered ‘no’ to both questions,

### Recommendations for the early phase of Covid-19 recovery

The first section of the survey addressed the immediate post discharge phase, that is, care or advice delivered in the home, in a step-down unit or in a rehabilitation hospital/ward that could be provided digitally or as a hard copy. Items that reached the threshold for recommendation for the early phase recovery programme are displayed in Table 1 (for complete data tables see online supplement). All proposed survey items were recommended for the early phase of Covid-19 recovery. There was overwhelming support for early post discharge from hospital phase of the recovery programme to advise patients about the management of fatigue (95% agreed or strongly agreed), breathlessness (94%), and mood disturbances (including symptoms of anxiety and depression) 92%. In recognition of the current UK community ‘lockdown’ there was clear agreement to provide support for coping with social isolation (91%). At this early stage in the recovery process there were less strong recommendations about cough management, delivery of an exercise programme or support for post-traumatic stress disorder but these comfortably exceeded the 70% threshold, at 81%, 80% and 78% respectively. Advice provided on a digital platform failed to reach the 70% threshold (59%), with 24.2% being neutral. This question provided the largest ‘neutral response’.

**Table 1:**
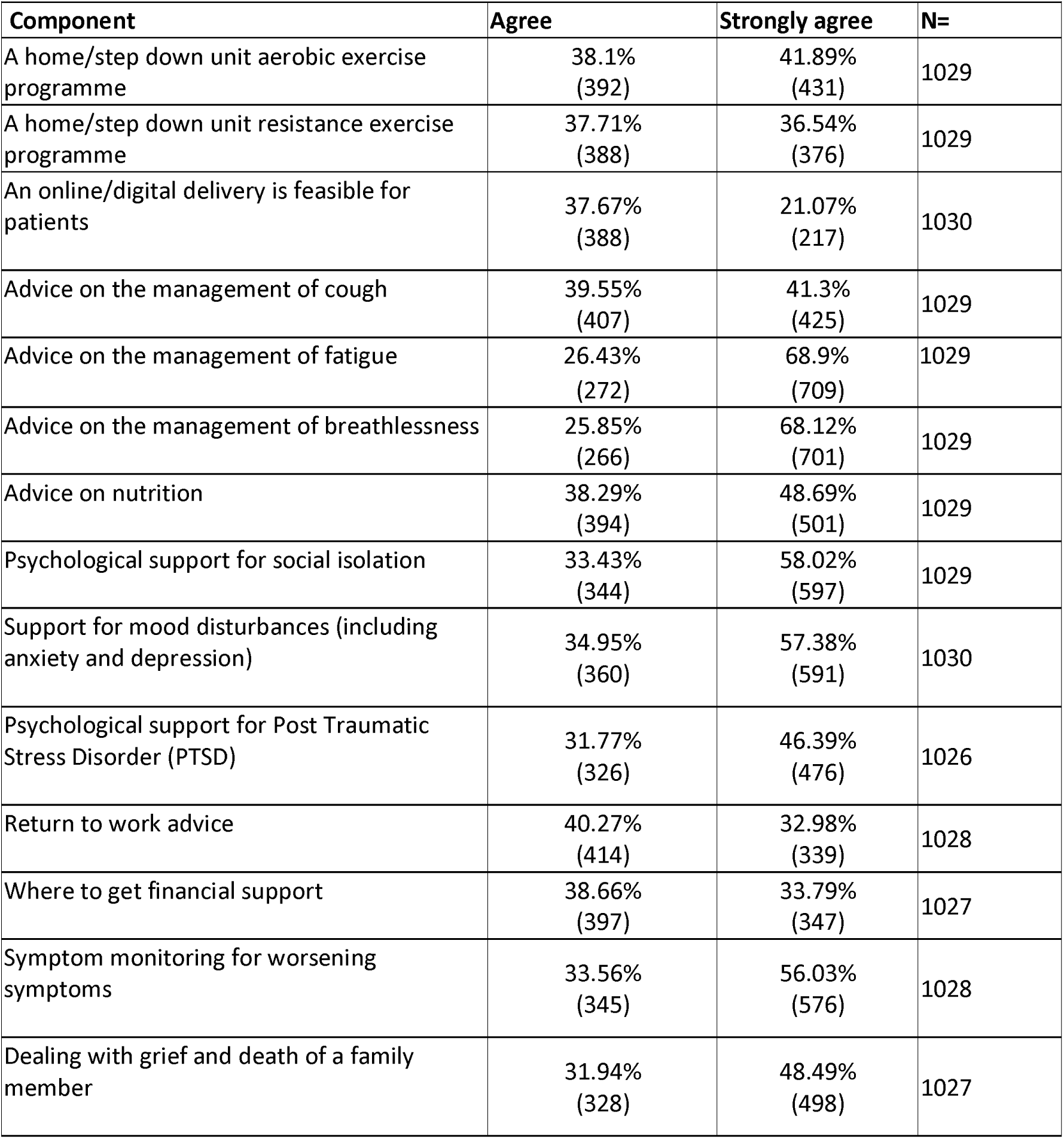
The essential components of an early phase recovery programme (first few weeks after discharge/episode)

### Recommendations for assessment at 6-8 weeks post hospital discharge

The essential components that reached consensus of an assessment at 6-8 weeks post episode/hospital (or step down unit) discharge are displayed in Table 2. There was strong support for the assessment of mood (93% strongly agreed or agreed), quality of life (92%) and fatigue (92%). The assessment of cough just reached the 70% threshold with 71% recommending assessment. Advice with respect to returning to work (73%) and financial support (72%) were not rated as highly although also exceeded the 70% threshold. The items which did not reach consensus were the need for a face to face assessment, assessment of exercise capacity/muscle strength and the need for a measure of lung function with 68%, 66% and 69% of the survey participants respectively recommending these factors as an important part of the assessment.

**Table 2:**
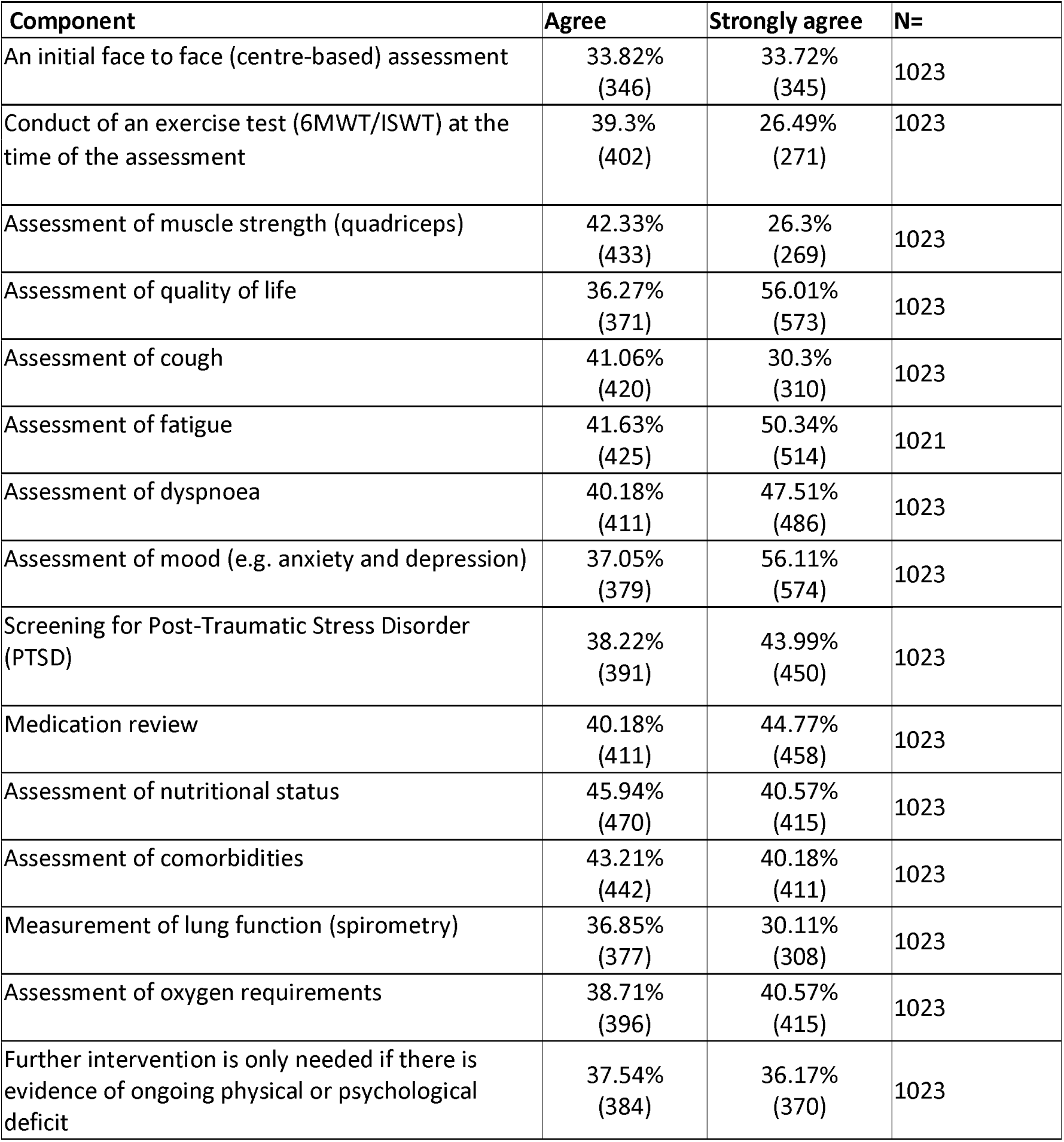
The essential components of an assessment at 6-8 weeks post hospital (step down unit) discharge

### Recommendations for the components of a rehabilitation recovery programme for Covid-19

The essential components which reached consensus for the later phase of recovery (6-8 weeks post discharge/episode and following the assessment outline above) are displayed in Table 3. The most frequently recommended items included advice on returning to usual exercise habits (93% either strongly agreeing or agreeing), 91% recommending advice on community exercise schemes, and given the ‘lockdown’ at the time of writing advice on community exercise schemes (once social isolation policy is relaxed) and advice for engaging in outdoors activities (once social isolation policy is relaxed) were highly rated (91 and 93% respectively) by respondents. Similarly exercise advice for home based aerobic and resistance programmes were highly rated (90% and 88% respectively). Symptom management was rated with advice on the management of fatigue and support for mood disturbances (including anxiety and depression) being equally strong recommended at 89%. Advice on the management of breathlessness was marginally less at 86%. Advice on the management of cough did not reach the 70% threshold at 6-8 weeks post discharge, with the largest number of respondents reporting to be ‘neutral’ for this question compared to any other question in this particular section (21%). The impact upon employment was also rated highly, advice on returning to usual employment (87%), where to get financial support advice (75%) and advice on returning to alternative employment (74%). Support for some unique aspects of Covid-19 and the current lockdown were also rated highly, psychological support for social isolation 84%, dealing with grief and death of a family member 80% and psychological support for Post-Traumatic Stress Disorder (PTSD) 80%. Assessment of lung function at 6 months post discharge was endorsed by 75% of respondents.

**Table 3:**
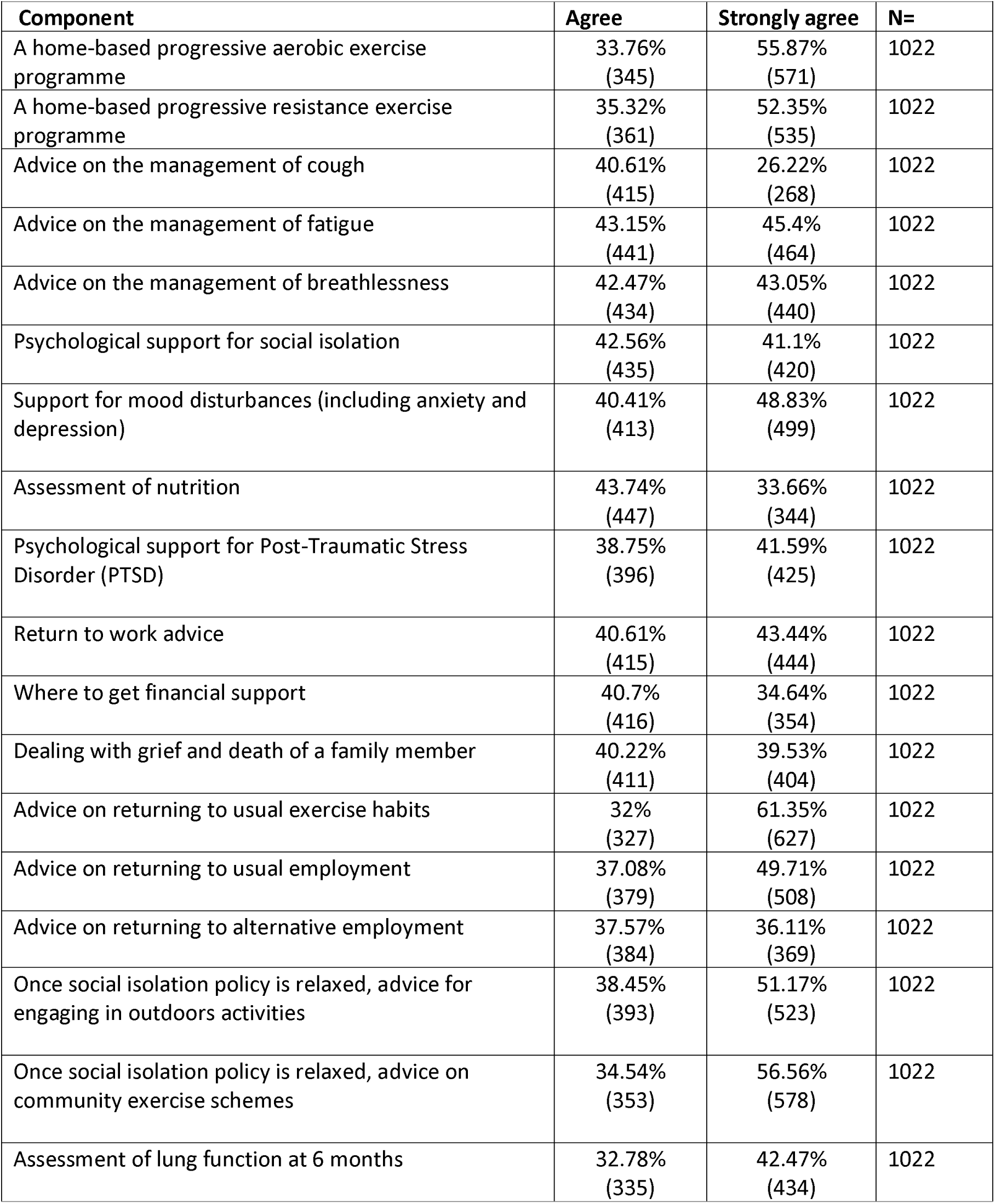
The essential components of a continued recovery programme beyond 6 weeks post hospital (step down unit) discharge

### Recommendations from the free text comments

A total of 341 free text comments were recorded and analysed. These informed seven themes and sub-themes. See Table 4 for illustrative quotes (further details in online supplement).

A large proportion of the results complimented the quantitative findings; however, additional service and treatment priorities were proposed. Firstly, respondents recognised that ‘A collaborative effort for rehabilitation development’ would be essential with input from experts in pulmonary/cardiac rehabilitation, nutrition, psychology, neurology, physiotherapy, respiratory medicine, occupational therapy and speech and language therapy, alongside recently published research from across the globe. Respondents felt there was a need to produce clear guidance for Covid-19 management, including this rehabilitation model, and there should be a campaign to promote Covid-19 rehabilitation to raise its profile amongst patients, carers and referrers and embed it within the Covid-19 recovery pathway.

Secondly, respondents recognised the uniqueness of this pandemic and therefore highlighted the importance of continued learning from Covid-19 for service development. It was recognised this would be an iterative process as services adapt to meet the new demands and service evaluations and research develop an evidence-based model.

Thirdly, alongside the early phase of recovery, suggestions for managing the acute phase were presented. Respondents highlighted the importance of assessing a patient’s physical and psychological wellbeing to inform personalised care plans. They also wanted to see a robust Covid-19 discharge bundle of self-management materials for both patients and caregivers.

The fourth theme comprised comments relating to the appropriate methods of rehabilitation delivery. Respondents felt this was an opportunity to adapt and improve current pulmonary rehabilitation models to meet the new demands and accommodate social distancing measures. For example, respondents suggested using pre and post outcome measures that could be assessed virtually (flexibility in assessment), using tele-rehabilitation with virtual group-based rehabilitation to maintain peer support. A personalised rehabilitation programme involving the assessment of patients’ care needs to inform a tailored rehabilitation plan from a menu of rehabilitation modules was proposed. There was debate about the timing of rehabilitation with some respondents leaning towards inpatient rehabilitation to minimise functional loss and others towards outpatient rehabilitation to allow time for immediate physical and psychological recovery. Access to rehabilitation was also acknowledged, with respondents highlighting the need for a clear referral pathway that healthcare professionals and patients can refer and re-refer to as necessary.

As a fifth theme, respondents proposed the necessary components for Covid-19 rehabilitation. There was acknowledgement of the effectiveness of current rehabilitation and holistic care pathways and therefore a desire not to reinvent the wheel, rather build on guidance from established rehabilitation models. Notably pulmonary rehabilitation, but other suggested models of care to consider and complement might include cardiac rehabilitation, neurorehabilitation, cognitive behavioural therapy, palliative rehabilitation, speech and language therapy, music therapy, yoga/tai chi, acupuncture, pastoral support and hydrotherapy. The majority of individual components recommended for a Covid-19 rehabilitation programme mirrored the quantitative findings, however, the following topics were also presented as care priorities: sputum clearance, frailty, pain, behaviour change, the impact of comorbidities, energy conservation, falls, inhaler technique, skin integrity, swallowing and voice care, cognitive functioning, inspiratory muscle training, caregiver support, signposting and peer support through group activities.

The sixth theme identified respondents wanting to see a team of specialist Covid-19 rehabilitation staff to deliver this new model. This is to include a multidisciplinary team who have specialist skills for this patient group. Additionally, respondents felt it was important to keep our staff physically safe, for example, by ensuring an appropriate supply of personal protective equipment (PPE) and the mental well-being of staff by monitoring and providing appropriate support when indicated to maintain the psychological health of the workforce.

Finally, respondents articulated the need for reassurance of financial support to ensure the robust development and delivery of this new rehabilitation model. They felt this support needed to be secured nationally to ensure equality and continuity of the service.

**Table 4:**
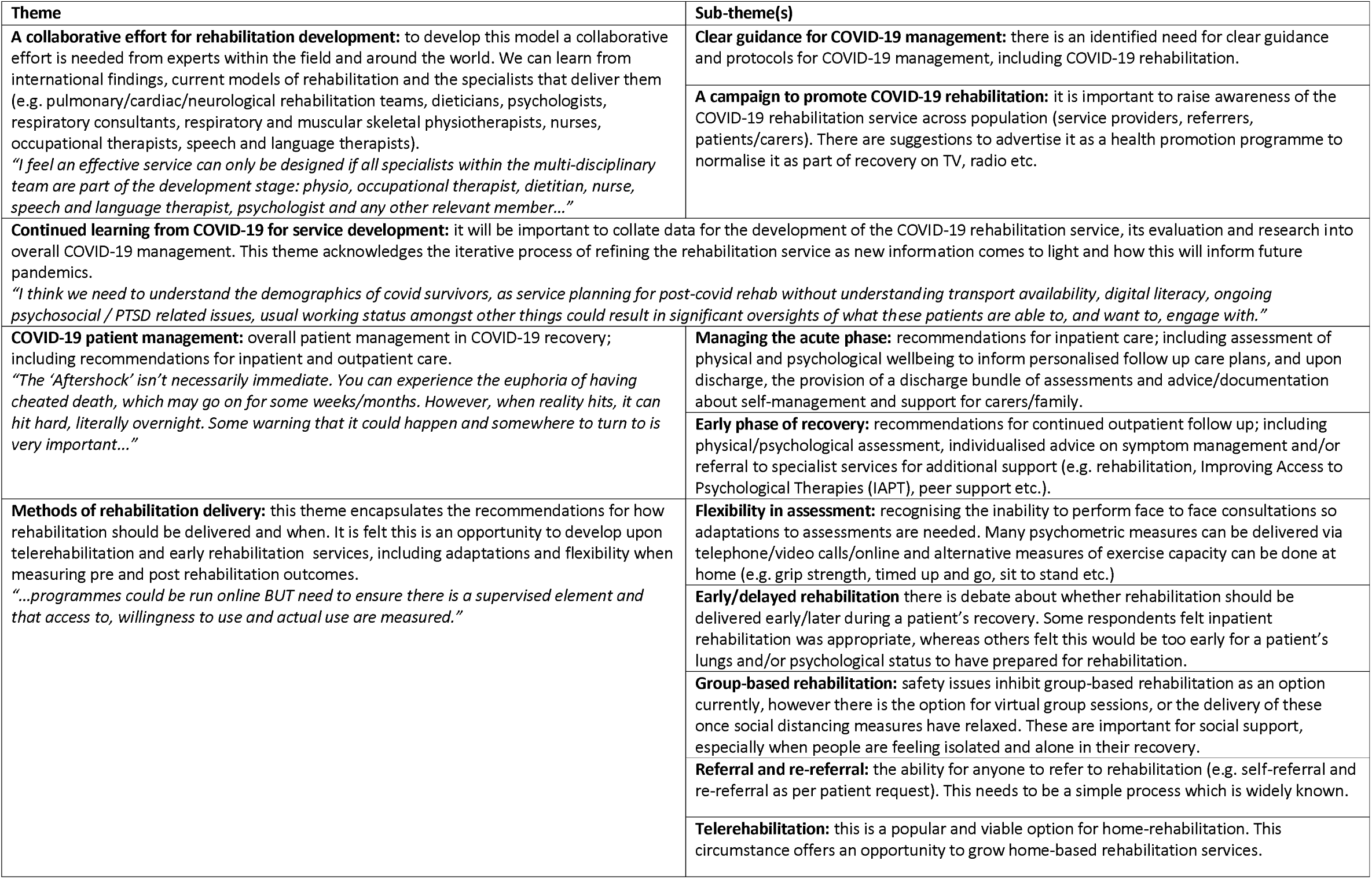
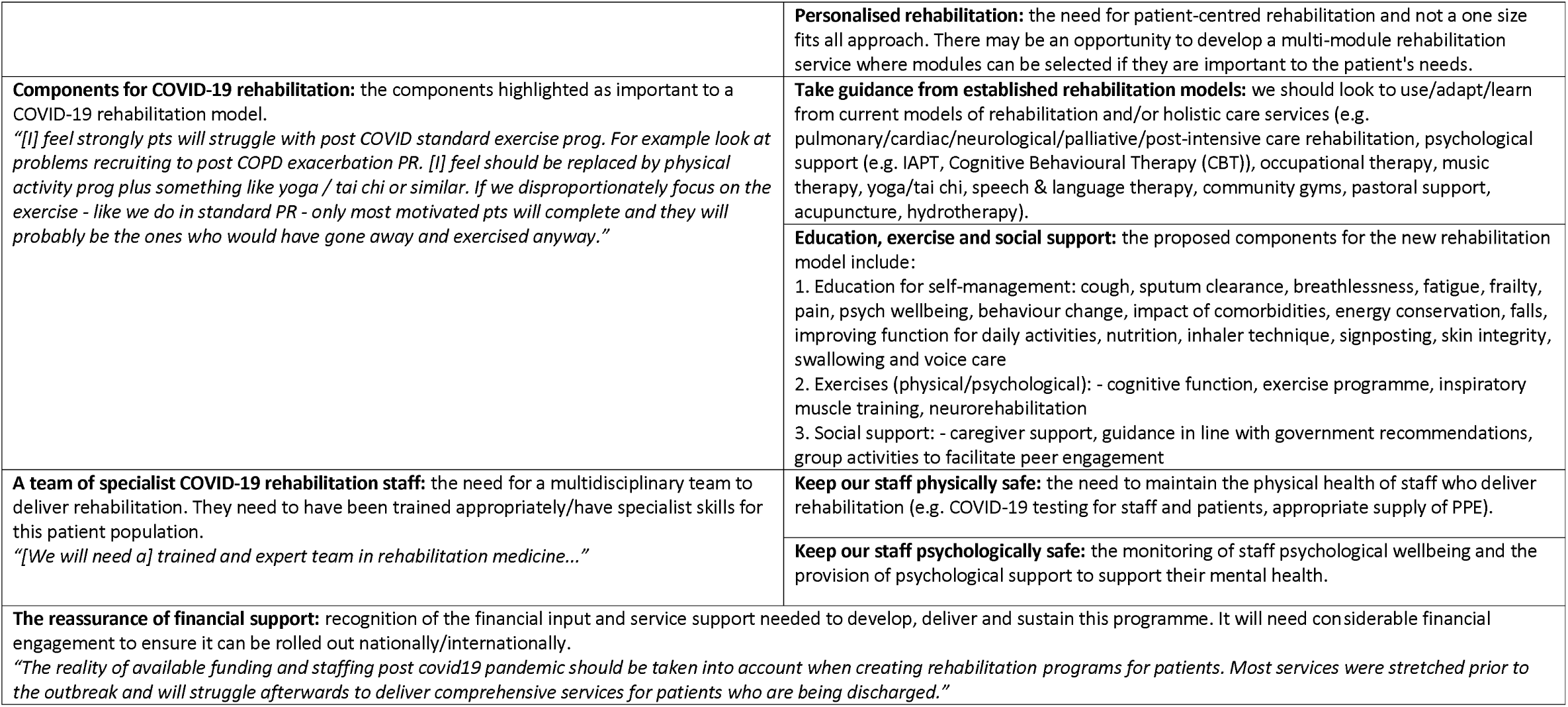
Generated themes and sub-themes from the survey’s free text comments

## Discussion

These data are the first from a comprehensive survey describing views from a large and diverse range of health-care professionals about the rehabilitation needs of the post Covid-19 population. Given the scale of response in such a short time period there is clearly a pressing need to develop a coherent recovery programme for people who are discharged from hospital after being infected with Covid-19. There was wide engagement with the health care community to support the development of the most appropriate package of rehabilitation, having secured the opinion of over 1,000 respondents from a wide variety of professional backgrounds and specialities. The survey identified the important components of the immediate post discharge phase, an assessment for a holistic rehabilitation intervention and the components of this intervention (Figure 1). The comments box allowed us to enrich the survey data and support the development of an appropriate recovery pathway for the post Covid-19 patient.

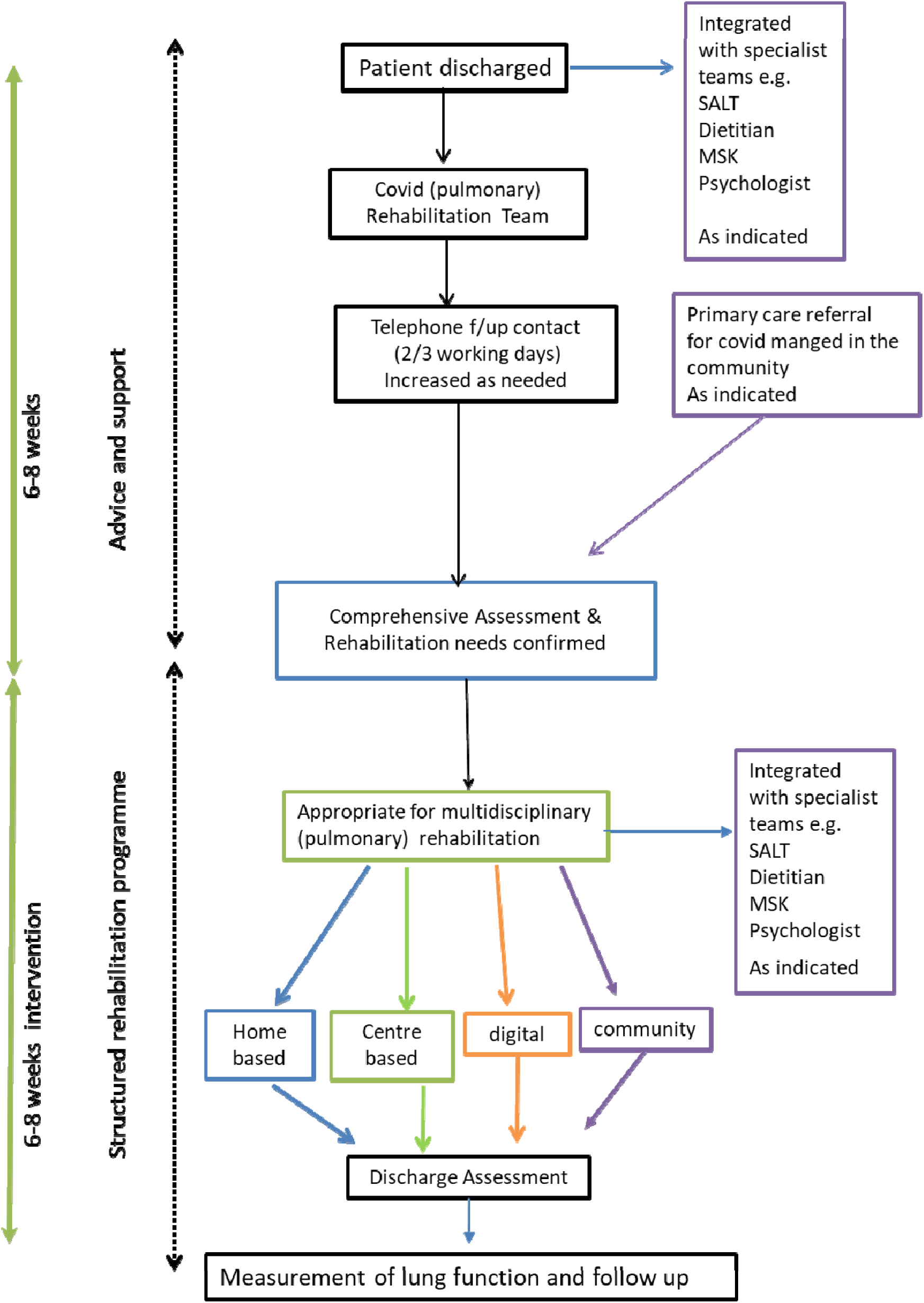

The predominant symptoms of Covid-19 are associated with the respiratory system (cough, breathlessness and fatigue) and the pulmonary rehabilitation model appears well placed to deliver the rehabilitation packages (with over 500 centres delivering pulmonary rehabilitation in the UK). Programmes currently deliver a personalised package of exercise and education, commonly extending over 6-8 weeks as a centre based, supervised package of care. Pulmonary rehabilitation is an interdisciplinary intervention that integrates a broad group of health care professionals including but not limited to physiotherapists, nurses, dietitians, pharmacists, psychologists, physicians, occupational therapists, exercise physiologists and graduates of the programme. As reported by respondents there is little appetite to ‘reinvent the wheel’ and develop a discreet single indication rehabilitation programme, rather there was a clear preference to adapt established pulmonary rehabilitation services to extend the scope to meet the needs of the post Covid-19 population. It was clear from the comments received that this needs to be collaborative and iterative as services become more experienced to meet the demands of this ‘new’ group. Many components recommended are already core components of a pulmonary rehabilitation pathway, assessment and programme, however the survey gave clear guidance of the additional components required to maximise impact, these included advice and support at the time of discharge. This is an important aspect of the cardiac rehabilitation pathway after discharge from cardiac revascularisation or myocardial infarction with routine telephone follow up [21]. The advice in the early stages focuses on symptom management and returning to normal (with a focus on gentle exercise and employment/financial issues). The assessment of the post Covid-19 patient at 6-8 weeks requires a much broader approach than commonly adopted by pulmonary programmes, specifically screening for PTSD and fatigue as a discreet symptom. PTSD is reported as a core outcome measure in the consensus statement for the follow up of ICU survivors [22], This report indicated that consensus was achieved for measures of mood, quality of life and PTSD, whilst exercise capacity and cognition almost reached consensus. It is beyond the scope of this survey to indicate the most appropriate outcome measures for the rehabilitation of the post Covid-19 population but there would seem a great deal of logic in combining the core measures of pulmonary rehabilitation with the outcomes recommended in the post ICU population. Interestingly the conduct of a face to face assessment, and performance of an exercise measure strength were recommended by 68%, and 66% respectively. These measures feature in all national and international guidance for PR and may reflect the current circumstances where face to face assessments are challenging. This is perhaps echoed in the free text comments with respect to service users and providers safety concerns. Additional comments from the survey identified the importance of measuring cognition, and importantly the need for integration with social care and Speech and Language Therapy. The timing and the modes of delivery was a discreet theme, and issues such as the feasibility of a face to face assessment and the need to be flexible in the current environment, with digital/telehealth solutions being highlighted as options. A sub-theme arose identifying the need for clear guidance. To date there is little guidance on the delivery of rehabilitation for this population. Previous international literature has described pulmonary rehabilitation services supporting recovery in other respiratory epidemics (SARS)[4] but the survey rightly reflects the need to collaborate with a much wider multidisciplinary team to offer the best service to patients post Covid-19.

This work highlights the real need for rehabilitating the post Covid-19 population and is strengthened by the large number of respondents. A limitation is that it did not consider the views of patients and the public; this is currently being undertaken by the British Lung Foundation[23]. These two surveys taken together should support guidance on the provision of rehabilitation services for the post Covid-19 patient.

It would seem that there is a real opportunity to develop a structured multidisciplinary rehabilitation programme that addresses the complex needs of the post Covid-19 population, alongside conventional pulmonary rehabilitation population. This would include those who had a period on ICU. The provision of post ICU rehabilitation although recommended[24] is poorly provided[25]. A legacy of this pandemic is the potential to raise the provision of post ICU care by integrating with established pulmonary rehabilitation services. However, it is important that capacity development is supported, as to not compromise the service for those who routinely access these programmes.

However, the more immediate challenge is to deliver a recovery pathway for those individuals who are being discharged now and for all those who have been discharged over the last few weeks with a diagnosis of Covid-19. We should use this survey data to inform service delivery and work collaboratively across specialties and professions to deliver a comprehensive recovery package for the Covid-19 population. Whilst of course retaining the high quality of service delivered to the usual case load of individuals with chronic respiratory disease.

## Conclusion

These data on over 1,000 respondents reflects the interest in the field of rehabilitation and the urgent need to adapt existing services to meet the complex set of needs that Covid-19 patients. Overall there was high level of agreement for the components of an early intervention, the elements of assessment and the components of the subsequent rehabilitation programme. This pandemic presents a real opportunity for truly collaborative working across disciplines and specialities and should be an immediate priority to mitigate the long term impact of Covid-19.

## Data Availability

No additional data are available.

## Authors contribution

SS, CEB GJ and JRH contributed to the conception and design of the study. LP, AB and SS analysed the data. SS and AB interpreted the data. SS and AB drafted the manuscript and all authors critically revised for significant intellectual content and insight. All authors had full access to all of the data and can take responsibility for the integrity and accuracy of data analysis. In addition, all authors gave final approval of the manuscript version for publication. SS and AB are responsible for the overall content as guarantors.

## Acknowledgements

The authors would like to thank the British Thoracic Society (particularly Sheila Edwards, Sally Welham and Kathryn Wilson) for supporting the concept of the survey, developing the online survey distributing to members and supporting the analysis. Furthermore, the authors would like to thank all the participants and those who promoted the survey to allow us to achieve the high response rate.

